# Alternative Approaches for Modelling COVID-19: High-Accuracy Low-Data Predictions

**DOI:** 10.1101/2020.07.22.20159731

**Authors:** Dewang Kumar Agarwal, Soham De, Ojas Shukla, Archit Checker, Apoorvi Mittal, Ankita Borah, Debayan Gupta

**Author notes:** Corresponding Authors: Dewang Kumar Agarwal; Debayan Gupta. These authors have equal contributions (Dewang Kumar Agarwal, Soham De, Ojas Shukla).

## Abstract

**Background:** Numerous models have tried to predict the spread of COVID-19. Many involve myriad assumptions and parameters which cannot be reliably calculated under current conditions. We describe machine-learning and curve-fitting based models using fewer assumptions and readily available data.

**Methods:** Instead of relying on highly parameterized models, we design and train multiple neural networks with data on a national and state level, from 9 COVID-19 affected countries, including Indian and US states and territories. Further, we use an array of curve-fitting techniques on government-reported numbers of COVID-19 infections and deaths, separately projecting and collating curves from multiple regions across the globe, at multiple levels of granularity, combining heavily-localized extrapolations to create accurate national predictions.

**Findings:** We achieve an *R*^2^ of 0·999 on average through the use of curve-fits and fine-tuned statistical learning methods on historical, global data. Using neural network implementations, we consistently predict the number of reported cases in 9 geographically- and demographically-varied countries and states with an accuracy of 99·53% for 14 days of forecast and 99·1% for 24 days of forecast.

**Interpretation:** We have shown that curve-fitting and machine-learning methods applied on reported COVID-19 data almost perfectly reproduce the results of far more complex and data-intensive epidemiological models. Using our methods, several other parameters may be established, such as the average detection rate of COVID-19. As an example, we find that the detection rate of cases in India (even with our most lenient estimates) is 2.38% - almost a fourth of the world average of 9% [1].

## 1 Introduction

On March 11, 2020, the World Health Organization (WHO) declared COVID-19 a pandemic [2], noting that there were “more than 118,000 cases in 114 countries, and 4,291 people have lost their lives.” Today, there are over 10 million confirmed cases. However, the availability and reliability of government-reported data remains questionable.

We employ a number of curve-fitting and machine-learning techniques to predict government reported numbers of infections in regions around the globe with high accuracy. In India, we note — with surprise — that this accuracy persisted through changes in testing policy, which suggests that these changes were not significant at the ground level.

Then, we describe a parameterized method of estimating the number of infections (including unreported infections) from the reported number of COVID-19 deaths. This uses demographics adjusted infection fatality ratio (IFR), disease progression, reporting rates, and growth statistics from recent literature and produces a count consistent with other, more complex, epidemiological models (INDSCI-SIM [3], MSEIRS [4], etc.).

These estimates may be used to compute other epidemiologically-significant variables, such as detection rate, which will hopefully be useful inputs to other models. While our predictions have been successful in predicting government-reported number in May and June (25th May to 16th June), we note that, with constantly evolving and updating numbers, the accuracy of our predictions can continuously improved by running our methods on the latest data available from reliable sources. **We have also applied our methods on current data (updated till 19th July, 2020) and report these predictions in Appendix F**. Our data, code, and models are available here: https://github.com/debayanLab/covidPredictions.

## 2 Methods

### 2.1 Data Sources

Infection statistics for all countries are collected from the official bodies of said countries via ourworldindata.org[5]. Case details and state-level statistics for India (till 9-May-2020) are derived from the Ministry of Health and Family Welfare (MoHFW) of India via collated datasets [6]. Health and medical data for Indian states are aggregated from Kaggle [7]. State-wise case progression for the United States was obtained from the New York Times [8] and [The COVID Tracking Project](https://covidtracking.com/data), updated daily. For the reported number of deaths in India, we use daily values provided by MoHFW, via ourworldindata.org [5]. Age structuring is derived from the 2011 Indian census projections [9].

### 2.2 Predicting Reported Cases

The pandemic strikes different regions around the globe at different times; so, mortality rate, R_0_, doubling time, etc. all vary. Therefore, we use curve-fitting and machine-learning models on national- and state-level data to predict government-reported numbers of total infections in multiple countries. We also study and use the case progression in other countries to predict progression in Indian states and districts, adjusting for around 24 medical and demographic variables through a multivariate and multi-output neural network. Ideally, we would perform projections for each hot-spot, and collate upwards, but that granularity of data is currently unavailable. We note that his method may be applied to any other country as well.

#### 2.2.1 Sigmoid Curve-Fitting

We use slope and rate-of-change of slope of the curve for total reported cases in India, the worst hit 10 states in the US, and European countries like France, Germany, and the UK; fitting and collating sigmoid curves from the district level for India and State level for the US, and country level for the European Countries adjusted for confounding factors. This is surprisingly accurate and is further confirmed by our machine-learning models.

##### Data Collection and Cleaning

###### India

Some data did not identify the district of a reported case. The state, however, was always identified. We assigned districts by weighted random assignment; the weight was the ratio of the cases in a district to that of its state. This was updated every time a case was added. We ignored districts with 0 cases. For all others, we started the day the first case was reported. Districts reporting an increase in infections for fewer than five days (across the period we were analyzing) were not subject to predictions using our logistic growth function. Instead, the prediction was simply the last reported total.

###### US States

Data was taken from the New York Times database [8]. From the date where the total number of reported cases was above 100 to 24th May was taken as the training data. From 25th May to 16th June was taken was testing data.

###### Europe

Data was taken from the Our World in Data database [5]. From the date where the total number of reported cases was above 100 to 24th May was taken as the training data. From 25th May to 16th June was taken was testing data.

The Gaussian mean (the peak value predicted by the normal distribution) and x_0_ (its x-coordinate) were calculated for each district/state/country (Appendix A). In case of India, predictions for each district were collated for the national prediction. Note that we do not currently account for regional differences in testing rate and migration.

#### 2.2.2 Machine-Learning

We observe that the curves in countries that have a significant number of cases like China and France closely resemble logistic growth. Further, similar trends in New York and Lombardy are visible as they progress through the pandemic.

Given these observations, we use machine-learning to fit the curve of Italy’s COVID-19 case progression (as Italy had almost finished its first wave and also had a lockdown) on Indian states. The model currently adjusts for around 24 medical and demographic variables; other useful variables were either difficult to represent, reduced model accuracy or unavailable.

Moreover, we use our global model to observe normalised time series data from China, Italy, South Korea, France, Spain, Germany, United States, the United Kingdom and all US states and territories to predict a global progression of the pandemic for the next 14 days.

##### Data Cleaning and Transformation

For our first model, daily counts were compiled to obtain a list of total cases and the ten Indian states with the most cases were listed. Similarly, COVID-19 cases in Italy were extracted and any missing values were forward-filled. Components (see Appendix B) from India’s census data that could directly affect the spread of cases were collated. All district-data were summed and appended to the state’s values. All three datasets were normalized within their respective axes and transformed into one-step-increment Long Short-Term-Memory (LSTM)-viable data.

For the second, global model, country and state-level time-series data was translated to start the day the region crossed 50 cases. This was then offset so the time series for all countries ended on the same day and any gap in the beginning was zero-padded. These values were then normalized along respective axes and reshaped to train our LSTM.

##### Training & Results

For both models, we employed the default sequential structure with the input layer of 200 Long-Short Term Memory (LSTM) nodes along with a rectified linear unit (ReLU) employed to introduce non-linearities, a repeat vector with dimensions of the number of outputs, a 200 LSTM nodes which returned sequences activated with ReLU, 100 time distributed dense nodes with ReLU activation and, finally, a single time distributed dense node.

An LSTM [10] is a Gated Recurrent Unit (GRU) which allows an RNN to learn long-term dependencies of sequential input data over extended periods, resulting in better time-series predictions.

For state-level predictions, primarily, the optimum input and output dimensions were achieved through loss comparison by sequential search of the variables. The maximum accuracy of correlation was obtained through a 24 time-step input and output. Therefore, 24 days of Italy’s cases along with census variables for each state, were utilized to output the forecasted number of cases for the coming 24-days in Indian states. Moreover, the model parameters were hyper-tuned using a random search method for 5 epochs for each of the 5 trails to achieve the the most accurate model without over-fitting data. The hyper-tuned, optimized state-wise model achieved an r-squared score of 0.991 for 24-day forecasts.

For our global predictions, we use a dense layer with 14 nodes to predict total cases for 9 countries and all US states and territories for the next 14 days. The input uses 24 prior time steps from time-series data from various countries/regions. To determine input length, we performed an empirical analysis of the loss obtained over different lengths, and chose the best one. To determine the best set of hyper-parameters, we used an automated tuner that optimizes for hyper-parameters in the specified space. The model predicted the progression of cases with a significantly high accuracy - an R-squared value of 0.995 on the test set.

### 2.3 Estimating the Number of Infections from Reported Deaths

When predicting active infections we assume that the actual number of infections are correlated more strongly to the reported number of fatalities than it is to the reported number of infections. From the reported number of deaths on a particular day, we back-calculate the expected number of infections on a previous day using disease progression reports and IFR estimates. While we do account for a time lag between the contraction of the virus and reporting of the death, this period is likely to differ among people and countries and involves a degree of uncertainty. Then, we use curve-fitting methods to extrapolate these numbers and make a prediction for the current day.

#### True Count of COVID-19 Deaths

To account for under-reporting of mortality statistics, we introduce a multiplier. Due to the strong dependence of this multiplier on the healthcare infrastructure of the country, it is separately evaluated for every country the method is applied on. For example, in India, the lower bound of this multiplier is 2, as countries reporting suspected as well as confirmed deaths establish a similar multiplier [11, 12]. The Million Deaths Study [13, 14] suggests that about 15% of all deaths are medically recorded in India. Accordingly, we assume 5 as a conservative upper bound in India. We apply similar methods for every country to determine these margins.

#### Disease Progression Statistics

It is reported that the mean incubation period for COVID-19 is around 5.1 days (95% CI 4.5–5.8 days) [15]. Estimates of the age-wise mean duration of onset of symptoms to death and delays in reporting of deaths were obtained from CDC [16] and weighed along the population demographics in the given country. This gives us an approximate period of 27 days from contracting the virus to reporting of death of a COVID-19 fatality in India and similar results on other countries as well. We also conduct several sensitivity analyses using other estimates [17, 18, 19] (see Appendix C).

#### Infection Fatality Ratio

There is evidence in literature that the IFR varies significantly with age-structure and quality of healthcare systems of a country. We use age-stratified IFR estimates from China [17] to calculate an age-weighted IFR estimate for a particular country (0.345% for India, based on the 2011 Census projections). We also consider other popular estimates [20, 17, 1, 21, 22, 23, 24] in our sensitivity analyses (see Appendix C).

#### Model for Disease Growth

We do not use any epidemiological models for the growth/spread of COVID-19. Instead, from our original assumption about reported deaths, we back-calculate time-shifted numbers of actual infections and then fit curves to extrapolate these numbers to the present day. In the initial stages of the pandemic, we found an exponential fit to be a closer fit, while with time, logistic curves better approximate these numbers.

We fit the following array of exponential and logistic curves and use the best fit among them for further analyses:

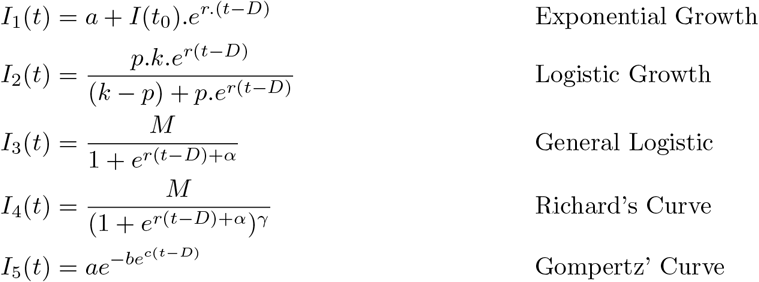

where D is mean time (in days) between contraction of infection and death, *r* is growth rate, *M* is population capacity; *α, γ, ρ* are curve fit parameters. Among these, the projections are most sensitive to the estimate of IFR used (see Appendix C). This method was applied on data from India, United States of America and certain European countries, both nationally and state-wise (wherever sufficient quality of data was available).

### 2.4 Calculating Present and Future Rate of Detection of COVID-19

#### 2.4.1 Estimated Rate of Detection at Present

The total infected number of cases, from 13th March till 5th June, were generated from reported deaths using disease parameters and logistic curve-fits. The total number of reported cases for the same time period were taken using ourworldindata.org [5]. Their average daily ratio gave the average present detection rate.

#### 2.4.2 Predicted Rate of Detection in Future

For each day after 6th June, the predicted number of reported cases was generated using our models in Section 2.2. Data from these models were divided by the corresponding predictions made daily by the four functions defined in Section 2.3. This gave us the daily predicted detection rate for a ten-day period from 6th June to 15th June (see Table 1).

**Table 1:**
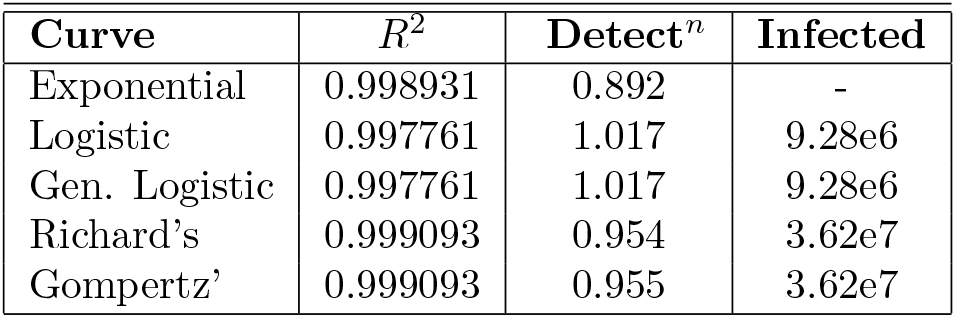
Curve Fitting Results (updated June 5)

| Curve | $R^2$ | Detect<sup>n</sup> | Infected |
| --- | --- | --- | --- |
| Exponential | 0.998931 | 0.892 | - |
| Logistic | 0.997761 | 1.017 | 9.28e6 |
| Gen. Logistic | 0.997761 | 1.017 | 9.28e6 |
| Richard's | 0.999093 | 0.954 | 3.62e7 |
| Gompertz' | 0.999093 | 0.955 | 3.62e7 |

## 3 Results

### 3.1 Total Number of Reported Cases

Let y_*i*_ be the ith day prediction including and after 25th May. Let n be the number of valid data points for the district. Let us define an 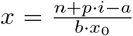. Then the prediction on the ith day for the district is given as

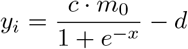

where m_0_ is the calculated Gaussian mean for each unit (district/state/country) and a, b, c, d, p are curve parameters. The parameters along with their average absolute percentage error between 25th march and 16th June for the ten worst hit US States, India, and three European countries have been summarized in Table 2 (see Appendix E). The state-level machine learning model performed with an R^2^ of 0.991 on the test set for 24 days of forecast, while the global model performed with a R^2^ of 0.995 on the test set.

**Table 2:** Comparison of parameters and average percentage error between 25th May and 16th June for the top 10 worst hit US States, India, and three European Countries using the Simple Functions model. The last column contains R^2^ values of Curve-Fits using the Richard’s Curve for the corresponding US States and Countries

| Region | a | b | c | d | p | Absolute % Error | Curve-Fit $R^2$ |
| --- | --- | --- | --- | --- | --- | --- | --- |
| New York | 0 | 1 | 1.3 | 133600 | 1.7 | 0.09 | 0.998 |
| New Jersey | 0 | 1 | 1.9 | -15000 | 1.6 | 0.20 | 0.999 |
| California | 50 | 1 | 10.5 | -2514000 | 1 | 0.84 | 0.998 |
| Illinois | -40 | 1 | 4.1 | 30000 | 1 | 0.47 | 0.999 |
| Massachusetts | 0 | 1 | 3 | 14000 | 4.5 | 0.70 | 0.999 |
| Texas | -120 | 1 | 9.8 | -4750 | 1.35 | 0.79 | 0.998 |
| Florida | 140 | 1 | 4.3 | -39500 | 2.5 | 1.42 | 0.998 |
| Pennsylvania | 0 | 1 | 20 | 360000 | 1.1 | 1.40 | 0.998 |
| Michigan | 0 | 1 | 5 | 93000 | 1.25 | 1.81 | 0.999 |
| Maryland | 50 | 1 | 4.7 | 4400 | 1.4 | 0.65 | 0.999 |
| India | 70 | 4.65 | 1180 | 5100 | 1 | 1.29 |  |
| France | 0 | 1 | 2 | -3300 | 1.65 | 0.42 | 0.998 |
| Germany | 0 | 1 | 4 | 67500 | 1.4 | 0.13 | 0.999 |
| UK | 70 | 1 | 9 | -94400 | 0.5 | 0.11 | 0.997 |

### 3.2 Actual Infections from Reported Deaths

We observe that Richard’s and Gompertz’ Curve gave us the best fit (R^2^ > 0.998) on the data from all the countries we ran our methods on. These include India, USA, United Kingdom, France and Germany. In general, all logistic curves are better approximates than exponential curves. While Richard’s Curve gives a slightly more accurate fit, Gompertz’ curve may also be used, as it may prevent overfitting due to the use of fewer parameters (see Table 3). As an example, we report the prediction for the actual number of infections in India, using both the kinds of curves discussed in figure 1. Results of application of the same methods on US state-wise data was similar and is noted in Appendix C.

**Table 3:** 10 day - Curve Fitting Projections by the 4 models in brackets and their corresponding Rate of Detection using predictions from the Simple Functions Model (starting from 06-June-2020).

| **Date** | **Prediction** | **Gompterz** | **Sigmoid** | **Richard's** | **Exponential** |
| --- | --- | --- | --- | --- | --- |
| 6th June | 246236 | 2.39<br>(10311477) | 3.05<br>(8062761) | 2.39<br>(10311103) | 1.81<br>(13588571) |
| 7th June | 255864 | 2.43<br>(10541724) | 3.15<br>(8132840) | 2.43<br>(10541318) | 1.81<br>(14103102) |
| 8th June | 265663 | 2.47<br>(10772926) | 3.24<br>(8199419) | 2.47<br>(10772488) | 1.82<br>(14636151) |
| 9th June | 275643 | 2.50<br>(11004999) | 3.34<br>(8262610) | 2.50<br>(11004527) | 1.81<br>(15188384) |
| 10th June | 285790 | 2.54<br>(11237857) | 3.43<br>(8322531) | 2.54<br>(11237350) | 1.81<br>(15760492) |
| 11th June | 296109 | 2.58<br>(11471417) | 3.53<br>(8379300) | 2.58<br>(11470873) | 1.81<br>(16353189) |
| 12th June | 306612 | 2.62<br>(11705595) | 3.64<br>(8433039) | 2.62<br>(11705013) | 1.81<br>(16967217) |
| 13th June | 317295 | 2.66<br>(11940308) | 3.74<br>(8483870) | 2.66<br>(11939686) | 1.80<br>(17603343) |
| 14th June | 328155 | 2.70<br>(12175476) | 3.85<br>(8531914) | 2.70<br>(12174812) | 1.80<br>(18262363) |
| 15th June | 339195 | 2.73<br>(12411016) | 3.95<br>(8577291) | 2.73<br>(12410309) | 1.79<br>(18945101) |

**Figure 1:**
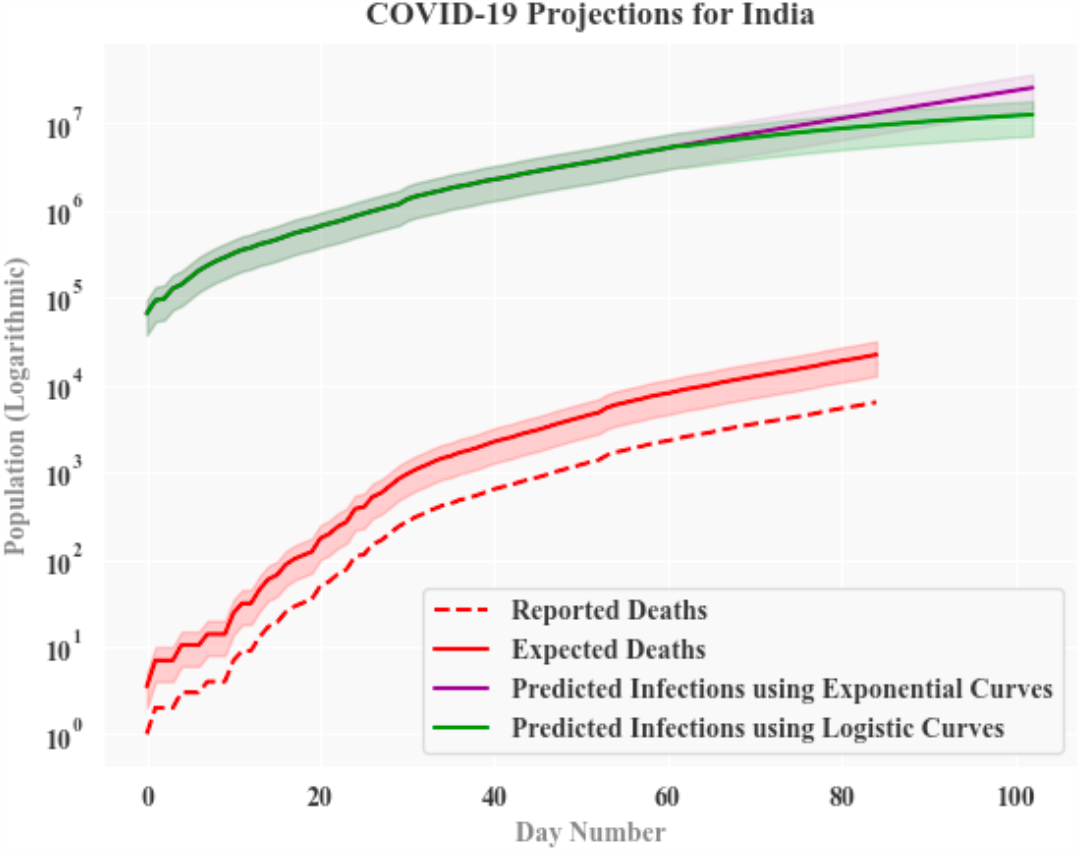
Actual Number of Infections, calculated from Reported Deaths in India. Green line denotes averaged prediction using all logistic curves. Purple line denotes predictions using exponential curves. Actual Number of Infections projected for a period of 18 days from 5th June

**Figure 2:**
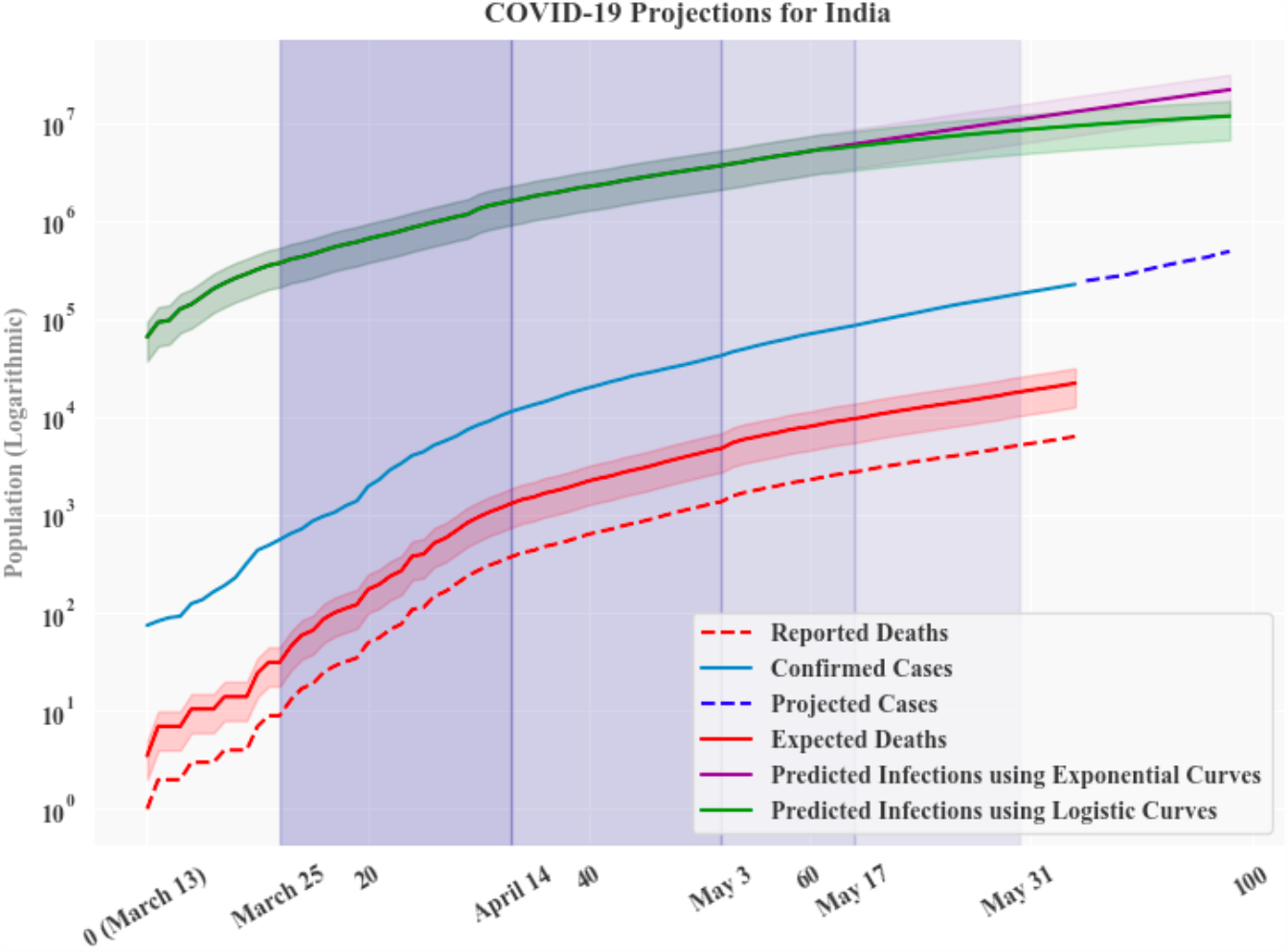
Actual Number of Infections, calculated from Reported Deaths (in green and purple) and Projection of Reported Number of Cases using curve fits (denoted in blue dotted lines). Period of projection: 18 days from 5th June

## 4 Discussion

Considering the neural network approach, our global prediction model utilizes trends from 9 countries that we chose due to the fact that these countries were among the ones with the most cases, and would follow the law of large numbers. We trained our model to predict this pattern on the basis of the growth in each of the selected countries, forecasting for the coming 14 days. We also use state-wise data from the United States and attain an R-Squared error of less than 1% on our test set. This model can be extrapolated to predict the growth of cases for any country, given we have a reasonable amount of data-points.Our state-level, curve-scaling, model was unable to provide a national prediction due to the lack of census data in some districts. Likewise, we believe that the error of the model could be reduced if we scaled using multiple countries, instead of only one (Italy).

Our simple functions model assumes that the average of the slope from the origin to each point on the curve is reasonably close to the slope of the origin to the Gaussian mean for the curve (see Appendix A). Low case numbers and slow daily increase might skew this average slope so we start training the model after 100 reported cases. Nevertheless, the function was able to predict the number of cases with an absolute percentage error of less than 1% (for 18 days) and less than 1.5% (over a period of 23 days). Given better testing statistics for each district or state, a more reliable district-wise or state-wise prediction could be scaled to the national level for multiple countries.Thee accuracy of this model suggests that it could be used to predict other parameters (such as active cases, number of deaths and number of cases healed) as well (see Appendix D). The prediction presented for these parameters are the number that the government or an authorised agency would report.

Possible ‘second-waves’ of COVID-19 in certain countries like the USA can also be indirectly accounted for using our methods. As most European countries are yet to show signs of a second wave, we have avoided any detailed description on the issue. However, while using our curve-scaling methods, the seconds waves may be treated as a superimposition of a sigmoid on an earlier sigmoid. Our Machine Learning based models should not require any further modification as LSTMs are generally superior at learning such trends in sequential data. If desired, additional ‘attention’ modules may be appended to the LSTMs to further improve accuracy.

While our methods may be applied on any country with a high number of reported COVID-19 cases, running them on data from India yeilds a detection rate of 2.9% in India, which falls in between the previously reported estimates of the same (1.45% [1] and 3.6% [21]). It may be noted that this rate is much lower than the world average of 9% [1] and is a genuine cause of concern. Recent serological survey results [25] show that 23.48% of people have been exposed to the virus. This is fairly consistent with our prediction of 29.7% as of 19th July. Other preliminary reports [26][27] of the ICMR serological survey, infections in hot-spots (e.g., Mumbai, Ahmedabad, etc.) are expected to be 100-200 times the number of reported cases (i.e detection rates of 0.5-1%), which is consistent with our estimates (we calculate an average detection rate of 0.688% for Maharashtra). These may serve as validation for our methods, which can safely be used on other countries, as we have demonstrated.

## Data Availability

The corresponding author confirms that he had full access to all data in this study and had final responsibility for submitting this manuscript for publication. All data has been taken from publicly available repositories. All the code used in the manuscript is provided in a public repository.

https://ourworldindata.org/coronavirus

https://github.com/nytimes/covid-19-data

https://api.covid19india.org/csv/latest/raw_data.csv

https://github.com/debayanLab/covidPredictions

## 5 Acknowledgment

The corresponding author confirms that he had full access to all data in this study and had final responsibility for submitting this manuscript for publication.

## References

[1] Sebastian Vollmer Christian Bommer. “Average detection rate of SARS-CoV-2 infections is estimated around nine percent”. In: (Mar. 2020). url: https://www.uni-goettingen.de/en/606540.html.

[2] URL: https://www.euro.who.int/en/health-topics/health-emergencies/coronavirus-covid-19/news/news/2020/3/who-announces-covid-19-outbreak-a-pandemic.

[3] Snehal Shekatkar & Bhalchandra Pujari & Mihir Arjunwadkar & Dhiraj Kumar Hazra & Pinaki Chaudhuri & Sitabhra Sinha & Gautam I Menon & Anupama Sharma and Vishwesha Guttal. INDSCI-SIM A state-level epidemiological model for India. Ongoing Study at https://indscicov.in/indscisim. 2020.

[4] Ashish Menon et al. “Modelling and simulation of COVID-19 propagation in a large population with specific reference to India”. In: (May 2020). doi: 10.1101/2020.04.30.20086306.

[5] Esteban Ortiz-Ospina Max Roser Hannah Ritchie and Joe Hasell. Coronavirus Pandemic (COVID-19). Published online at OurWorldInData.org. Retrieved from ‘https://ourworldindata.org/coronavirus. 2020.

[6] covid19india.org. url: https://api.covid19india.org/csv/latest/raw_data.csv.

[7] Indian State Census Data. 2020. url: https://www.kaggle.com/webaccess/india-census-yearly-data.

[8] The New York Times. An ongoing repository of data on coronavirus cases and deaths in the U.S. The New York Times. Available at: https://github.com/nytimes/covid-19-data. 2020.

[9] Registrar General and Census Commissioner of India. Census of India 2011: Provisional Population Totals. Ministry of Home Affairs, New Delhi, India. Available at: http://www.censusindia.gov. 2011.

[10] Sepp Hochreiter and Jürgen Schmidhuber. “Long Short-term Memory.” In: Neural computation (1997). doi: 1735-80.10.1162/neco.1997.9.8.1735.

[11] Tracking covid-19 excess deaths across countries, The Economist. Retrieved from https://www.economist.com/graphic-detail/2020/04/16/tracking-covid-19-excess-deaths-across-countries. 2020.

[12] Wall Street Journal. Most countries fail to capture extent of COVID-19 deaths. 2020. url: https://www.wsj.com/articles/most-countries-fail-to-capture-extent-of-covid-19-deaths.

[13] Rukmini S. In India, most deaths go unregistered. How reliable is its Covid-19 mortality data? 2020. url: https://scroll.in/article/961081/in-india-most-deaths-go-unregistered-how-reliable-is-its-covid-19-mortality-data.

[14] Erica Westly. Global health: One million deaths. 2013. url: https://www.nature.com/news/global-health-one-million-deaths-1.14269.

[15] Stephen Lauer et al. “The Incubation Period of Coronavirus Disease 2019 (COVID-19) From Publicly Reported Confirmed Cases: Estimation and Application”. In: Annals of internal medicine 172 (Mar. 2020). doi: 10.7326/M20-0504.

[16] A. and Barnard L. Wilson N. and Kvalsvig and Baker M. G. “Case-Fatality Risk Estimates for COVID-19 Calculated by Using a Lag Time for Fatality”. In: Emerging Infectious Diseases Journal (June 2020). doi: https://dx.doi.org/10.3201/eid2606.200320.

[17] Robert Verity et al. “Estimates of the severity of coronavirus disease 2019: a model-based analysis”. In: The Lancet Infectious Diseases (Mar. 2020). doi: 10.1016/s1473-3099(20)30243-7. url: https://doi.org/10.1016/s1473-3099(20)30243-7.

[18] Shankar R. Gupta Sourendu. “Estimating the number of COVID-19 infections in Indian hot-spots using fatality data”. In: 162.5 (Sept. 2020), pp. 479–486. doi: https://arxiv.org/pdf/2004.04025.pdf. URL: https://arxiv.org/pdf/2004.04025.pdf.

[19] Jun Chena and Tangkai Qia et al. “Clinical progression of patients with COVID-19 in Shanghai, China”. In: Journal of Infection (Mar. 2020). doi: 10.1016/j.jinf.2020.03.004. url: https://doi.org/10.1016/j.jinf.2020.03.004.

[20] Gideon Meyerowitz-Katz and Lea Merone. “A systematic review and meta-analysis of published research data on COVID-19 infection-fatality rates”. In: (May 2020). doi: https://doi.org/10.1101/2020.05.03.20089854.

[21] Debraj Chakraborty Siuli Mukhopadhyay. “Estimation of undetected COVID-19 infections in India”. In: (MedRxiv Preprint) (May 2020). doi: 10.1101/2020.04.20.20072892. url: https://doi.org/10.1101/2020.04.20.20072892.

[22] John P.A. Ioannidis. “The infection fatality rate of COVID-19 inferred from seroprevalence data”. In: (MedRxiv Preprint) (Mar. 2020). doi: 10.1101/2020.05.13.20101253. url: https://doi.org/10.1101/2020.05.13.20101253.

[23] Russell Timothy W et al. “Estimating the infection and case fatality ratio for coronavirus disease (COVID-19) using age-adjusted data from the outbreak on the Diamond Princess cruise ship”. In: Euro Surveillance (Feb. 2020). doi: 10.2807/1560-7917. url: https://doi.org/10.2807/1560-7917.

[24] Giulio A. De Leoa Richard E. Grewellea. “Estimating the global infection fatality rate of covid-19”. In: (MedRxiv Preprint) (May 2020).

[25] url: https://indianexpress.com/article/cities/delhi/delhi-serological-survey-covid-19-icmr-6516208/

[26] S. Staff. Covid-19: ICMR Rejects Reports That Said Over 15% People In Hotspots Infected, Says Study Not Final. Retrieved from https://scroll.in/latest/964183/covid-19-icmr-rejects-reports-that-said-over-15-people-in-hotspots-infected-says-study-not-final. 2020.

[27] ICMR’s Sero-Surveillance Study Reveals Only 0.73% of the Sample Population Infected with COVID-19. Retrieved from ddnews.gov.in/national/icmr\OT1\textquoterigh sero-surveillance-study-reveals-only-073-sample-population-infected-covid-19. 2020.

[28] Johns Hopkins Coronavirus (COVID-19) Dataset. url: https://gisanddata.maps.arcgis.com/apps/opsdashboard/index.html#/bda7594740fd40299423467b48e9ecf.

[29] Gupta and Shankar. Estimating the number of COVID-19 infections in Indian hot-spots using fatality data. Apr. 2020. url: https://arxiv.org/abs/2004.04025.

